# Cross-ancestry and phenome-wide associations of cancer-specific polygenic risk scores

**DOI:** 10.1101/2025.05.15.25327704

**Authors:** Louise Wang, Heena Desai, Craig Teerlink, Oliver Clark, Kathryn M. Pridgen, VA Million Veteran Program, Jennifer Lee, Kyong-Mi Chang, Rachel L. Kember, Marijana Vujkovic, Julie A. Lynch, Kara N. Maxwell

**Affiliations:** VA Connecticut Healthcare System, West Haven, CT; Section of Digestive Diseases, Department of Medicine, Yale School of Medicine, New Haven, CT; Division of Gastroenterology and Hepatology, Department of Medicine, Perelman School of Medicine, University of Pennsylvania, Philadelphia, PA; Corporal Michael J. Crescenz VA Medical Center, Philadelphia, PA; Division of Hematology/Oncology, Department of Medicine, Perelman School of Medicine, University of Pennsylvania, Philadelphia, PA; Division of Epidemiology, Department of Internal Medicine, University of Utah School of Medicine, Salt Lake City, UT; Veterans Affairs Informatics and Computing Infrastructure, Salt Lake City, UT; Division of Internal Medicine, Pennsylvania Hospital, University of Pennsylvania, Philadelphia, PA; VA Palo Alto Healthcare System, Palo Alto, CA; Division of Endocrinology, Department of Internal Medicine, Stanford School of Medicine, Palo Alto, CA; Department of Psychiatry, Perelman School of Medicine, University of Pennsylvania, Philadelphia, PA; Division of Translational Medicine and Human Genetics Department of Genetics, Perelman School of Medicine, University of Pennsylvania, Philadelphia, PA; Department of Genetics, Perelman School of Medicine, University of Pennsylvania, Philadelphia, PA; Abramson Cancer Center, Perelman School of Medicine, University of Pennsylvania, Philadelphia, PA

**Keywords:** cancer, polygenic risk score, pleiotropy, cross-cancer associations, polygenic risk

## Abstract

**Background:** Genome-wide association studies (GWAS) have identified many common variants associated at low effect sizes with various cancers. Summing the effects of these variants into polygenic risk scores (PRS) can improve cancer risk prediction. However, these PRS have largely been analyzed in cohorts restricted to individuals of European descent, and research into potential cross-cancer and cross-phenotype pleiotropic associations of cancer-specific PRS is limited.

**Methods:** Using logistic regression models, we tested the association of 13 cancer-specific PRS (bladder, colorectal, esophageal, glioma, lung, melanoma, oral cavity, pancreatic, renal, thyroid, breast, prostate, and testicular) with curated phenotypes representing the same 13 cancers and 340 cancer and cardiometabolic phecodes in 560,287 individuals (114,255 African ancestry, AFR and 446,032 European ancestry, EUR) from the Million Veteran Program (MVP), a large, ancestrally-diverse biobank. Logistic regression models were stratified by ancestry and used age, principal components, and cancer-specific PRS as independent variables.

**Results:** All 13 cancer-specific PRS were significantly associated (p<0.002) with their respective cancers among EUR individuals (OR 1.05 to 1.70). Among AFR individuals, cancer-specific PRS were significantly associated with their respective cancers for five of 13 cancers (bladder, breast in females, colorectal, prostate, and thyroid, OR 1.19 to 1.48). In cancer-cancer pleiotropy studies, only the renal cancer-specific PRS was significantly associated with skin cancer (OR=1.04, P=4.5×10^−06^) among EUR individuals. PheWAS demonstrated five positive pleotropic associations with cardiometabolic conditions (thyroid cancer PRS with thyroid goiter, oral cancer PRS with type 1 diabetes, ophthalmic and renal diabetic complications, and hypothyroidism) and two negative associations (oral and lung cancer PRS separately with coronary artery disease). There were no significant cancer-phecode associations among AFR individuals.

**Conclusions:** We confirm the validity of 13 cancer-specific PRS on predicting their corresponding cancer in MVP, observing stronger associations per cancer among EUR versus AFR individuals. In contrast to PRS of other chronic diseases, our study shows that the majority of cancer-related PRS are highly specific and pleiotropic associations with other cancers and cardiometabolic traits are relatively uncommon.

## Introduction

Susceptibility to cancer results from a combination of environmental factors and genetic risk. Genome-wide association studies (GWAS) have identified common genetic variants associated with many cancers, but, for the majority, the associated risk with each single variant is low^1,2^. Polygenic risk scores (PRS), defined as a risk-weighted sum of multiple to genome wide genetic alleles, separate individuals’ risk of a disease across a broader range of absolute risk than a single variant. PRS may be helpful in risk prediction and optimizing population-based screening for many diseases, including cancers^3,4^. Previous studies have examined the predictive performance of multiple cancer-specific PRS in large biobanks^5–7^. However, these studies were limited by either small sample size or cohort demographics such as tertiary care settings^6^, which may not reflect true population-based risk, and/or predominantly individuals of European genetic ancestry (EUR)^5,7^. The lack of ancestral diversity in studies examining the predictive performance of PRS is a significant research gap.^8,9^ We and others have previously shown that while PRS may remain statistically significantly associated with a disease, they generally show inferior performance in risk prediction in populations of African ancestry (AFR) compared to EUR populations, likely due to the fact that the majority of PRS uses weights derived from EUR-predominant GWAS^2,8–13^.

Pleiotropy, when one gene or variant is associated with multiple phenotypes, has been identified among several cancer susceptibility variants, and shared heritability is well known for many variants across multiple cancers.^7,14^ Further, given that cancer and cardiovascular diseases share a number of environmental and clinical risk factors^15,16^, there may be shared genetic pathways of oncogenesis and development of cardiometabolic disease. Phenome-wide association studies (PheWAS) offer the opportunity to explore the pleiotropic effects of known risk variants across the entire phenome and may increase the understanding of underlying shared biological pathways between diseases^17^. Identification of these shared variants may facilitate improved genetic testing for multiple cancers or repurposing of treatment therapies between disease entities^18,19^.

In this study, we aimed to measure the performance of 13 cancer-specific PRS for cancer prediction in EUR and AFR individuals and to evaluate the phenome-wide associations of cancer PRS with other cancers and cardiometabolic phenotypes in the diverse Million Veteran Program (MVP), the largest electronic health record (EHR)-linked megabiobank in the world.

## Methods

### Study population

We performed a retrospective, case-control study of participants in MVP, the largest EHR-linked multi-ethnic genetic biorepository of United States citizens^20,21^. Since 2011, Veterans receiving healthcare within the Veterans Health Administration (VHA) have been recruited from more than 75 VA Medical Centers. Participants in this study were identified as having AFR or EUR ancestry as defined by harmonized ancestry and race/ethnicity (HARE), which incorporates self-reported and genetically-inferred ancestry using a machine learning-derived algorithm^22,23^.

Informed consent was obtained from all participants to provide blood for genomic analysis and access to their full EHR data within the VA prior to and after enrollment, including inpatient International Classification of Diseases (ICD)-9 or ICD-10 diagnosis codes. This retrospective cohort study used data analyzed as part of an MVP research study protocol that was approved by the US Department of Veterans Affairs (VA) Central Institutional Review Board. All participants provided written informed consent and Health Insurance Portability and Accountability Act authorization (what we used to submit on med archive).

### Genotyping data

This study used the MVP Release 4 genotyping data which has data for EUR (n=446,032) and AFR (n=114,255) individuals on a customized Affymetrix Axiom biobank array (the MVP 1.0 Genotyping Array), which contains >753,000 genetic markers that have been imputed to TopMed Reference Panel version 2^24^. Details on the quality control, imputation, and ancestry determination have been described previously^20,22^.

### Cancer phenotypes

We identified individuals diagnosed with 13 cancer groupings (bladder, colorectal, esophageal, glioma, lung, melanoma, oral cavity, pancreatic, renal, thyroid, breast [male and female separately], prostate, testicular) from the VA Corporate Data Warehouse (CDW) who had at least one inpatient or outpatient International Classification of Diseases (ICD)-9/10 diagnosis code (**Table S1**). We also defined a more stringent cancer phenotype that required at least four separate instances of the ICD-9/10 diagnosis code for a particular cancer in a sensitivity analysis. We defined controls as individuals with no ICD-9/10 codes for any invasive cancer, benign, in situ, or secondary neoplasms (**Table S1**). Given the low proportion of female participants in the biobank, we exclusively included males as cancer/control cases except when we analyzed the breast cancer PRS separately in females and males.

### PRS construction

We obtained GWAS summary statistics for each of the 13 cancers using the GWAS catalog for that cancer as previously described^6,13^ (**Table S2**), except for breast and prostate cancer for which we used previously established PRS^25,26^. We selected SNPs based on a p-value threshold (P<1.0×10^−06^) and pruned for linkage disequilibrium (LD) at r^2^ < 0.1 using the EUR 1000 Genome database reference panel in PLINK 1.9^27^. Using PLINK 1.9, we generated individual level cancer-specific PRS by summing the LD-pruned SNPs weighted by their corresponding effect sizes. For the previously validated prostate cancer PRS, we calculated PRS using effect sizes from the GWAS of EUR and AFR individuals separately.

### PRS performance of cancer-specific PRS

For each cancer PRS, we performed logistic regression analysis stratified by HARE on our outcome (i.e., cancer cases) to determine the odds ratio (OR) of cancer per standard deviation of the PRS, controlling for age and the first 10 principal components (PCs) as covariates. The significance level for this analysis was set at a Bonferroni correction of 0.002 due to 28 tests. We calculated the additional discriminatory performance of including PRS in a risk assessment versus using only age and the first 10 PCs using under the curve (AUC) via the pROC package in R 4.3.1.

### PheWAS to understand secondary cancer and cardiometabolic phenotypic associations

Using the R PheWAS package^28^, we performed PheWAS for each cancer PRS that was significantly associated with its cancer-specific ICD code analysis (n=13 PRS in EUR, n=5 PRS in AFR). We then identified the 19 phecodes that correspond most closely to the ICD codes used for the 13 cancers above and six additional cancer types (liver and bile duct, stomach, Hogkin’s disease, non-Hodgkin’s lymphoma, leukemia, myeloproliferative disease, **Table S3**). We also selected 340 phecodes from the PheWAS catalog in the circulatory system category (394-459) and the endocrine/metabolic category (240-278)^17^. We identified significant associations at a Bonferroni corrected p-value threshold p<1.39×10^−4^ and the overall Phewas significant threshold of p<1×10^−6^. These PheWAS were performed with phecodes as the outcome variables, with cancer PRS as the independent variable, and age and the first 10 PCs as covariates in multiple logistic regression models.

## Results

### Baseline characteristics

Among the 560,287 MVP participants, 163,849 (29%) had at least one of the studied cancer phenotypes (22,164 AFR males, 95,649 EUR males, 1,591 AFR females, 4,703 EUR females). The median age of cases at enrollment was 68 years (median age 65 in AFR, 69 in EUR). Among AFR individuals, the most common cancers included prostate (14,704, 66.3%), lung (2,888, 13.0%), colorectal (2,578, 11.6%), renal (2,009, 9.1%), and bladder (1,357, 6.1%) cancer. Among EUR individuals, the most common cancers included prostate (44,255, 46.27%), lung (14,355, 15.0%), melanoma (12,973, 13.6%), bladder (11,748, 12.3%) and colorectal (10,936, 11.4%). We compared cancer cases to cancer-free controls (51,030 AFR and 180,328 EUR, median age at enrollment 62 years old) (**Table 1, Table S4**).

**Table 1:** Cancer cases and controls within MVP.

| Cancer | HARE-European |  |  | HARE-African |  |  |
| --- | --- | --- | --- | --- | --- | --- |
|  | Controls | n-1 | n-4 | Controls | n-1 | n-4 |
| ***Cancers tested in males only*** |  |  |  |  |  |  |
| Bladder | 180328 | 11748 | 7964 | 51030 | 1357 | 735 |
| Colorectal | 180328 | 10936 | 6681 | 51030 | 2578 | 1475 |
| Esophageal | 180328 | 2595 | 1556 | 51030 | 324 | 163 |
| Glioma | 180328 | 1807 | 710 | 51030 | 375 | 110 |
| Lung | 180328 | 14355 | 9620 | 51030 | 2888 | 1927 |
| Melanoma | 180328 | 12973 | 5766 | 51030 | 215 | 48 |
| Oral Cavity | 180328 | 5371 | 3298 | 51030 | 899 | 475 |
| Pancreatic | 180328 | 2501 | 1334 | 51030 | 630 | 332 |
| Renal | 180328 | 7606 | 4609 | 51030 | 2009 | 1251 |
| Thyroid | 180328 | 2210 | 1488 | 51030 | 391 | 246 |
| Prostate | 180328 | 44255 | 33729 | 51030 | 14704 | 12317 |
| Testicular | 180328 | 1489 | 710 | 51030 | 222 | 67 |
| ***Cancers tested in males and females*** |  |  |  |  |  |  |
| Breast (males) | 180328 | 479 | 229 | 51030 | 156 | 68 |
| Breast (females) | 20705 | 2021 |  | 12071 | 825 |  |

n-1: Number of individuals with $\geq 1$ ICD9/10 code for the cancer
n-4: Number of individuals with $\geq 4$ ICD9/10 code for the cancer

### Cancer-specific PRS performance

Among EUR individuals, all 13 cancer-specific PRS were significantly associated with their respective cancers identified by ICD code grouping at a Bonferroni corrected p-value threshold of p<0.002 (**Figure 1a, Table S5**). The PRS with the strongest associations (expressed as odds ratio per standard deviation of PRS) with their respective cancer types were those for prostate cancer (OR 1.70, 95% CI 1.68–1.72, p<1e^-300^), breast cancer in females (OR 1.40, 95% CI 1.33–1.47, p=6.3e^-44^), melanoma (OR 1.35, 95% CI 1.32–1.37, p=1.6e^-248^) and colorectal cancer (OR 1.27, 95% CI 1.24–1.29, p=8.2e^-123^).

**Figure 1:**
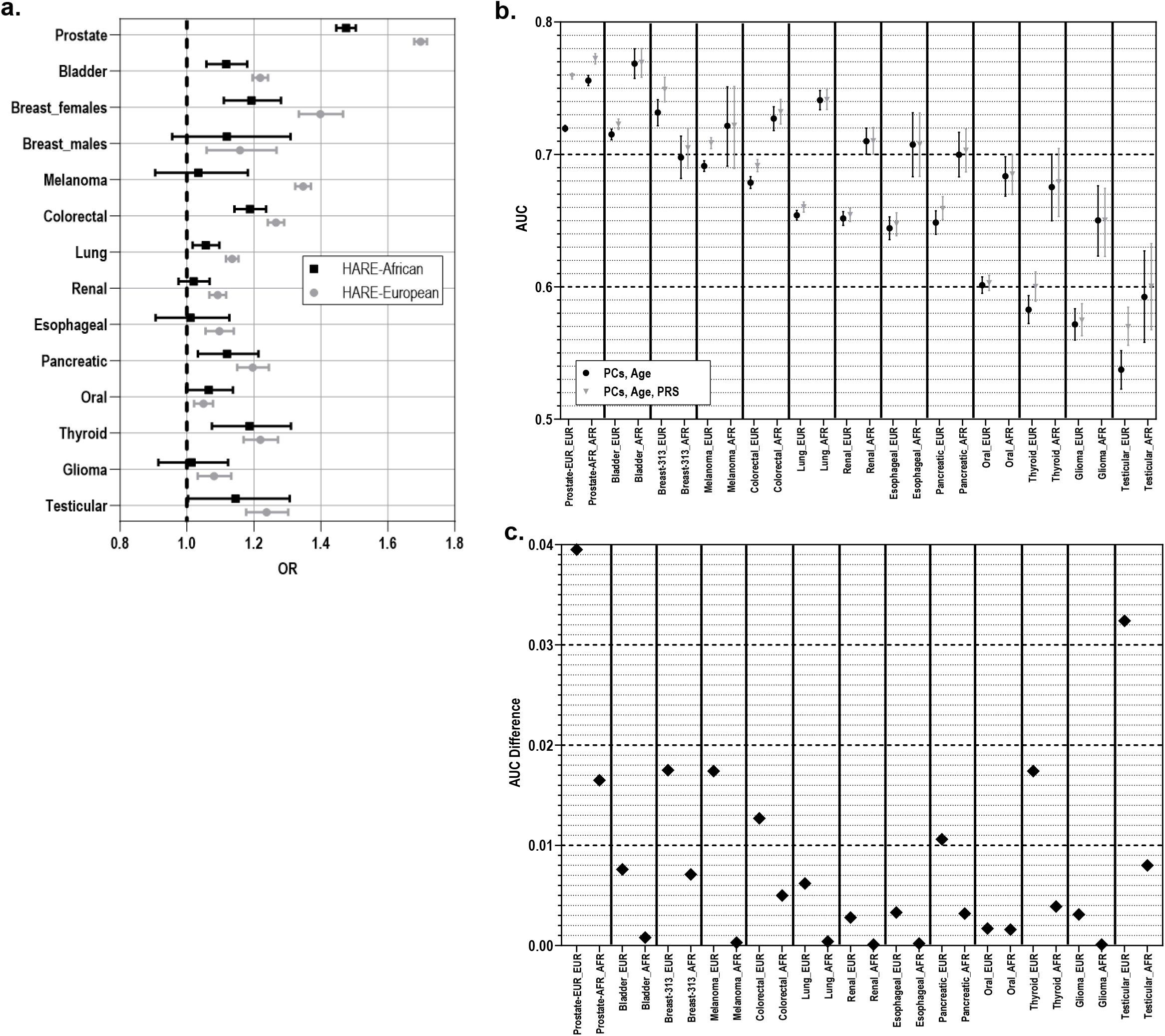
Performance of Cancer PRS in Million Veterans Program. **a**. Odds ratios per standard deviation of PRS for association of each cancer PRS with its corresponding cancer in EUR and AFR individuals where cases were defined as having at least one instance of the corresponding cancer ICD9/10 code. **b**. AUCs for models with and without PRS for EUR and AFR individuals where cases were defined as having at least one instance of the corresponding cancer ICD9/10 code. **c**. AUC difference with and without PRS for EUR and AFR individuals where cases were defined as having at least one instance of the corresponding cancer ICD9/10 code.

Among AFR individuals, the following five cancer-specific PRS were significantly associated (p<0.002) with their respective cancers: prostate cancer (OR 1.48, 95% CI 1.45– 1.50, p<1e^-300^), breast cancer in females (OR 1.19, 95% CI 1.11–1.28, p=1.4e^-6^), colorectal cancer (OR 1.19, 95% CI 1.14–1.24, p=2.3e^-17^), thyroid cancer (OR 1.19, 95% CI 1.07–1.31, p=7.1e^-4^), and bladder cancer (OR 1.12, 95% CI 1.06–1.18, p=6.7e^-5^). (**Figure 1a, Table S6**). Additional nominal associations (p<0.05) were seen for cancers of the lung (OR 1.05, 95%CI 1.02-1.10, p=4.9e^-3^), pancreas (OR 1.12, 95%CI 1.03-1.21, p=6.0e^-3^), and testis (OR 1.15, 95%CI 1.00-1.31, p=0.04).

For each cancer type, we evaluated the additional discriminatory performance of an adding PRS to models containing age and the first 10 PCs. Among EUR individuals, the PRS significantly increased the discriminatory performance beyond age and the first 10 PCs alone for cancers of the prostate (ΔAUC: 0.040, p<1e^-15^), testis (ΔAUC: 0.032, p=3.3e^-6^), breast in females (ΔAUC: 0.018, p=3.8e^-12^), melanoma (ΔAUC: 0.017, p<1e^-15^), thyroid (ΔAUC: 0.017, p=3.3e^-6^), colon and rectum (ΔAUC: 0.013, p<1e^-15^), pancreas (ΔAUC: 0.011, p=6.4e^-7^), bladder (ΔAUC: 0.008, p<1e^-15^), lung (ΔAUC: 0.006, p<1e^-15^), and kidney (ΔAUC: 0.003, p=6.5e^-6^) (**Figure 1b-c, Table S7**). Among AFR individuals, only the prostate and colorectal cancer PRS significantly increased the discriminatory performance beyond age and the first 10 PCs (prostate, ΔAUC: 0.017, p<1e^-15^; colorectal, ΔAUC: 0.005, p=5.5e^-5)^, **Figure 1b-c, Table S8)**. In all cancers, the PRS increased discriminatory ability beyond age and PCs by a greater amount in EUR compared to AFR as measured by ΔAUC (**Table S7, Table S8**).

We conducted a sensitivity analysis using a more stringent cancer phenotype (≥4 encounters for ICD-9 or ICD-10 diagnosis code) for all cancer-PRS pairs except breast cancer in males, which we excluded due to insufficient number of cases (**Table 1**). Except for breast cancer in females, gliomas, and oral cavity cancer, ten cancer PRS remained significantly associated with their respective cancers in EUR individuals (**Figure S1a, Table S9)**. In AFR individuals, three of five (prostate, colorectal, and bladder cancer) PRS remained significant (**Figure S1a, Table S10)**. Among both individuals of AFR and EUR ancestry, the association was stronger for most cancer PRS in both genetic ancestries. The discriminatory ability of the PRS addition to existing model with age, and 10 PCs was similar between the stringent and standard case phenotype definitions (**Figure S1b-c, Tables S11-S12**).

### Cancer PRS with cancer phecode and cross-cancer associations

To investigate whether any cancer-specific PRS predicted risk of other cancers, we performed PheWAS with the 13 cancer-specific PRS across 19 phecodes for common cancers (**Table S3, Table S13**). In EUR individuals, the bladder, breast, colorectal, lung, melanoma, pancreatic, prostate, testicular and thyroid PRS were significantly associated with their respective cancer phecode (**Figure 2a, Table S13**) and esophageal, glioma and renal showed nominal associations (p<0.05). With the exception of bladder cancer, the OR of cancer PRS-phecode pairs was marginally higher (2-18% increase) than their respective OR of cancer PRS-ICD pairs (**Table S14**). In AFR individuals, only the prostate cancer PRS was significantly associated with its respective phecode; bladder, colorectal, lung, and testicular were nominally associated (**Figure 2b, Table S13**). For cross-cancer associations, we found an association between renal cancer PRS with melanomas of the skin (OR 1.04, p: 4.53 × 10^−6^). Additional nominal associations between PRS and cross-cancer phenotypes based on phecodes can be seen in **Figure 2a**. Among AFR individuals, no cross-cancer associations with phenome-wide significance were observed (**Figure 2b**). We noted nominal associations between esophageal cancer and the bladder, breast, and oral cancer PRS.

**Figure 2:**
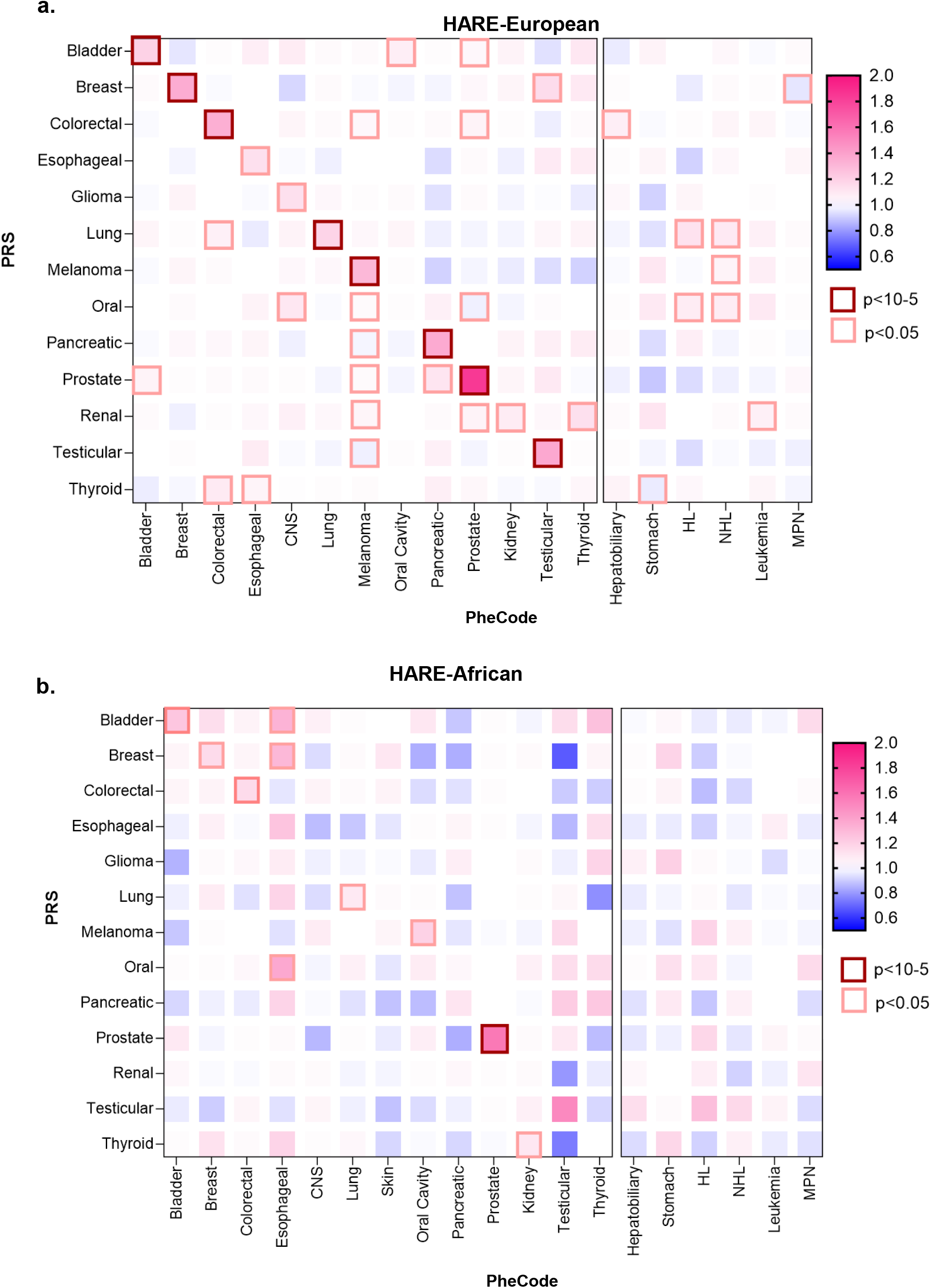
Association of Cancer PRS with PheCodes in Million Veteran Program. **a**. Odds ratio and p-value in a Phewas testing for association of each cancer PRS with 19 cancer phecodes among EUR individuals. **b**. Odds ratio and p-value in a Phewas testing for association of each cancer PRS with 19 cancer phecodes among AFR individuals.

### Cardiometabolic and phenome-wide associations

The PheWAS revealed significant associations between cancer PRS and phecodes within circulatory system and endocrine/metabolic categories only among EUR individuals (**Table S15, Figures S2-S14**). We found five positive associations: thyroid cancer PRS with nontoxic nodular goiter, and oral cavity PRS with type 1 diabetes, two diabetic complications (ophthalmic and renal), and hypothyroidism. Additionally, both the oral and lung cancer PRS had a significant negative association with coronary artery disease manifestations (e.g., coronary atherosclerosis and ischemic heart disease).

## Discussion

Using the largest multi-ancestry biobank linked to an electronic health record, MVP, we evaluated the predictive performance of 13 cancer-specific PRS and assessed for cross-cancer and cardiometabolic pleiotropy. We found significant associations for all 13 cancer-specific PRS in EUR individuals, but significant associations for only five PRS despite a large sample set of AFR individuals (n=114,255). PRS significantly increased discriminatory ability over age and genetic principal components in ten cancers in EUR, and two cancers in AFR. In pleiotropy studies, we only identified one cross-cancer association and five cancer-cardiometabolic phenotype associations. Overall, most cancer PRS were specific and had few cross-cancer or cardiometabolic associations.

Previous studies have shown poorer predictive performance of PRS among individuals of non-EUR ancestry^6,29^, largely because many GWAS have been performed among majority EUR individuals^8,30^. In our study, the prostate cancer PRS, the only PRS with base summary statistics exclusively from AFR individuals, had the best predictive performance among AFR individuals. Among both ancestries, prostate cancer PRS provided the greatest improvement in model discrimination beyond age and first 10 PCs, which may be related to the high genetic hereditability of prostate cancer^31,32^.

The only significant cross-cancer association we found was between the renal cancer PRS and melanoma, consistent with prior studies showing a bidirectional association between renal-cell carcinoma and melanoma^33^. We did not confirm any of the previously reported cancer PRS pleiotropic associations reported in a UK Biobank study^7^. However, individuals in the Million Veteran Program may also have military exposures that could contribute to their cancer risk^34^, and future studies should incorporate their unique military exposures into their cancer risk profile. Single SNP level cross-cancer associations have been identified^35^; however, these were not investigated in the current study.

Cancer and cardiac disease share several environmental risk factors and may develop due to aberrations in similar biological pathways. However, competing mortality may mask associations or suggest inverse associations between these two diseases. In fact, prior studies have observed an inverse association between polygenic risk for coronary artery disease and incident breast^36,37^ and lung cancer^36^. We found no positive associations between any cancer PRS and any cardiac phecode. We did identify a negative association between oral and lung cancer PRS and coronary artery disease manifestations. Further work is needed to determine if this association results from shared genetic architecture in opposite effects, genetic predisposition of cancers resulting in subsequent behavioral modifications (e.g., smoking cessation or improved dietary patterns)^38^, competing risks of death prior to a cancer diagnosis, protective treatment effects for CAD against cancer, or ascertainment bias with the use of diagnosis codes.

With regards to associations of cancer PRS with metabolic phenotypes, our study identified a statistically significant association between thyroid cancer PRS and nontoxic nodular goiter, likely either from increased detection of nontoxic nodular goiter during ultrasound testing^39^ or overlapping common variant germline genetics between thyroid cancer and goiter.

We also identified a positive association between the oral cavity PRS and autoimmune conditions such as type 1 diabetes and hypothyroidism. One possible shared mechanism is Epstein-Barr virus, which shuttles between epithelial and B cells and has been shown to increase risk for both oral cavity cancer^40^ and autoimmune conditions^41^.

Our study has multiple strengths. First, our study included over 560,000 individuals and nearly 164,000 cancer cases from a diverse, population-based VA cohort spanning over 75 enrollment sites across the US, simulating both the random risk of a general population but also the at-risk ages of cancer. Second, MVP is a diverse cohort that provides valuable non-European individual-level data for PRS. We also analyzed multiple phenotype definitions of the various cancers with similar findings for each, which served as internal validation.

Our study has several limitations. First, we identified prevalent not incident cancer cases within MVP. Additionally, while we included age and first 10 PCs in our analysis, we did not include family history, body mass index (BMI), or other medical comorbidities which could result in a confounding bias due to shared genetic and environmental factors with PRS. Thirdly, the performance of PRS was still better in the EUR ancestry group than in the AFR ancestry group, though this effect was mitigated in the prostate cancer PRS which was calculated using base data from AFR individuals. As more GWAS are performed within cohorts including AFR individuals, we will likely see improvements in PRS performance among this population. Lastly, because the full summary statistics were not available for each cancer, we were limited in our methodological approaches for PRS to statistical thresholding.

In summary, we confirmed the validity of cancer-specific PRS in predicting corresponding cancer within MVP, with stronger cancer PRS associations among EUR individuals than AFR individuals. In contrast to PRS of other chronic diseases, most cancer-related PRS are highly specific and pleiotropic associations with other cancers and cardiometabolic traits are relatively rare.

## Supporting information

Supplemental Acknowledgements

## Acknowledgements

We gratefully acknowledge the Veterans who participated in the Million Veteran Program. This research is based on data from the Million Veteran Program, Office of Research and Development, Veterans Health Administration, and was supported by MVP003/028 as well as award I01-BX003362. The authors acknowledge the support of the MVP Core Team (**Supplement**). This work was supported using resources and facilities of the Department of Veterans Affairs (VA) Informatics and Computing Infrastructure (VINCI), including data analytics conducted by its Precision Medicine research team, which is funded under the research priority to Put VA Data to Work for Veterans (VA ORD 24-D4V-02). This publication does not reflect views of the Department of Veterans Affairs or the United States government.

## Data Availability Statement

It is not possible for the authors to directly share the individual-level data that were obtained from the MVP due to constraints stipulated in the informed consent. Anyone wishing to gain access to this data should inquire directly to MVP at. The data generated from our analyses are included in the manuscript main text, tables, and figures and online Supplementary Materials (available online). The summary data for MVP are available through dbGAP, accession number phs001672.

