## Supplemental Figures for "Cross-ancestry and phenome-wide associations of cancer-specific polygenic risk scores"

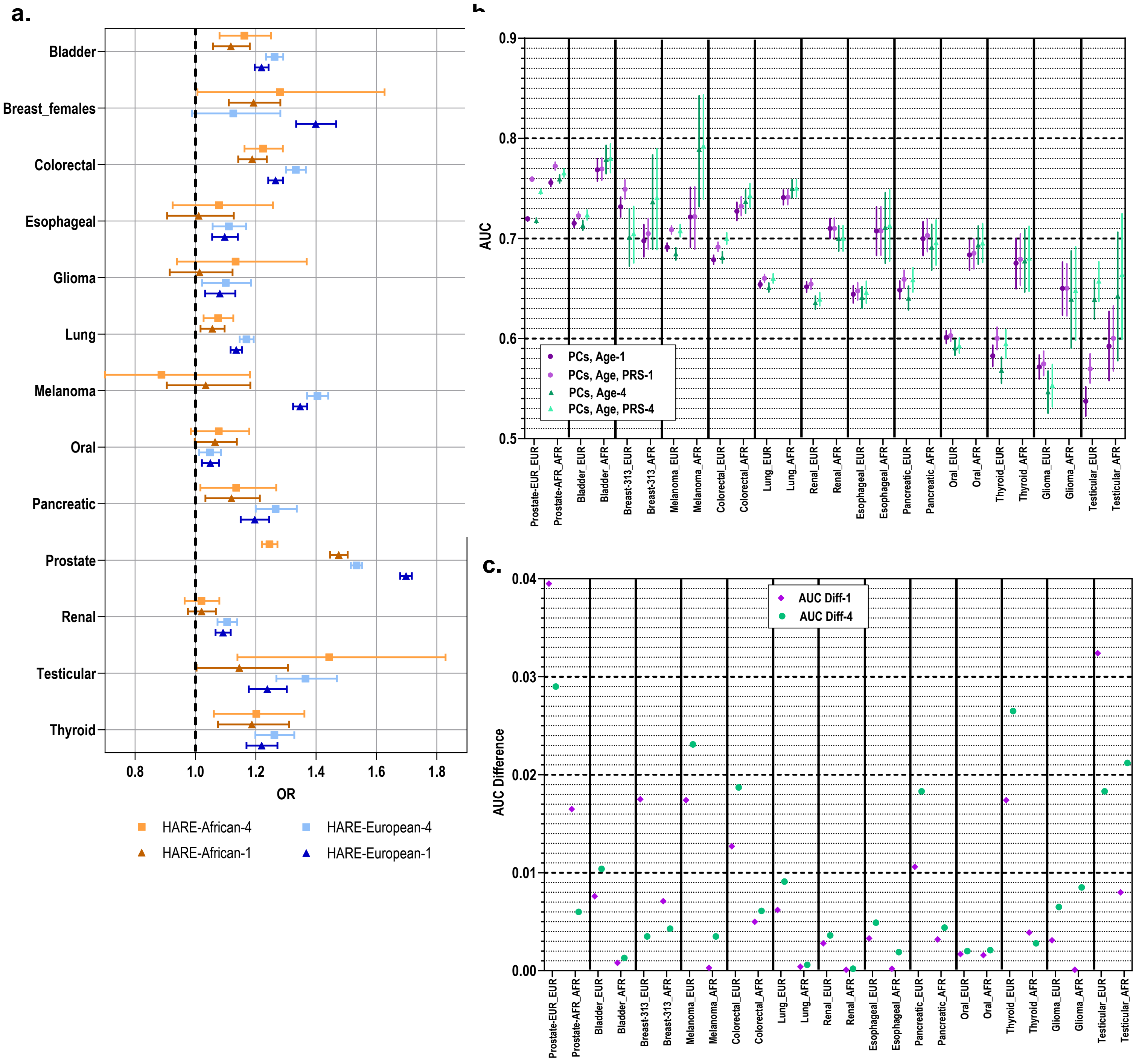


**Supplemental Figure 1: Performance of Cancer PRS in Million Veterans Program. a.** Odds ratios for association of each cancer PRS with its corresponding cancer per standard deviation of PRS in HARE-European and HARE-African individuals where cases were defined as having at least one versus at least four instances of the corresponding cancer ICD9/10 code. **b.** AUCs for models with and without PRS for HARE-European (EUR) and HARE-African (AFR) individuals where cases were defined as having at least one or at least four instances of the corresponding cancer ICD9/10 code. **c.** AUC difference with and without PRS for HARE-European (EUR) and HARE-African (AFR) individuals where cases were defined as having at least one or at least four instances of the corresponding cancer ICD9/10 code.

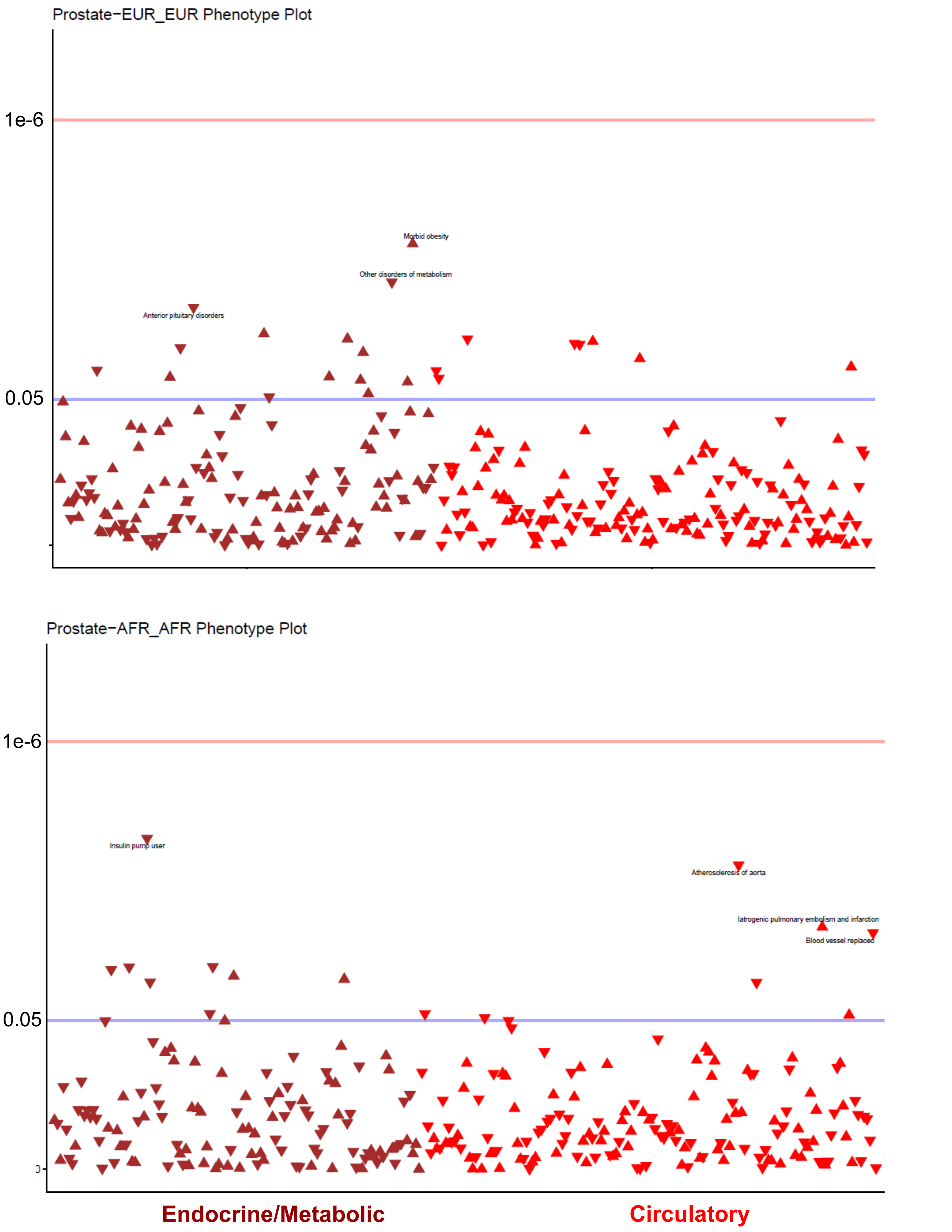


**Supplemental Figure 2: Association of prostate cancer PRS with cardiometabolic phenotypes.** Phewas to examine the association of Prostate cancer PRS with circulatory and metabolic/endocrine phecodes in HARE-European and HARE-African individuals.

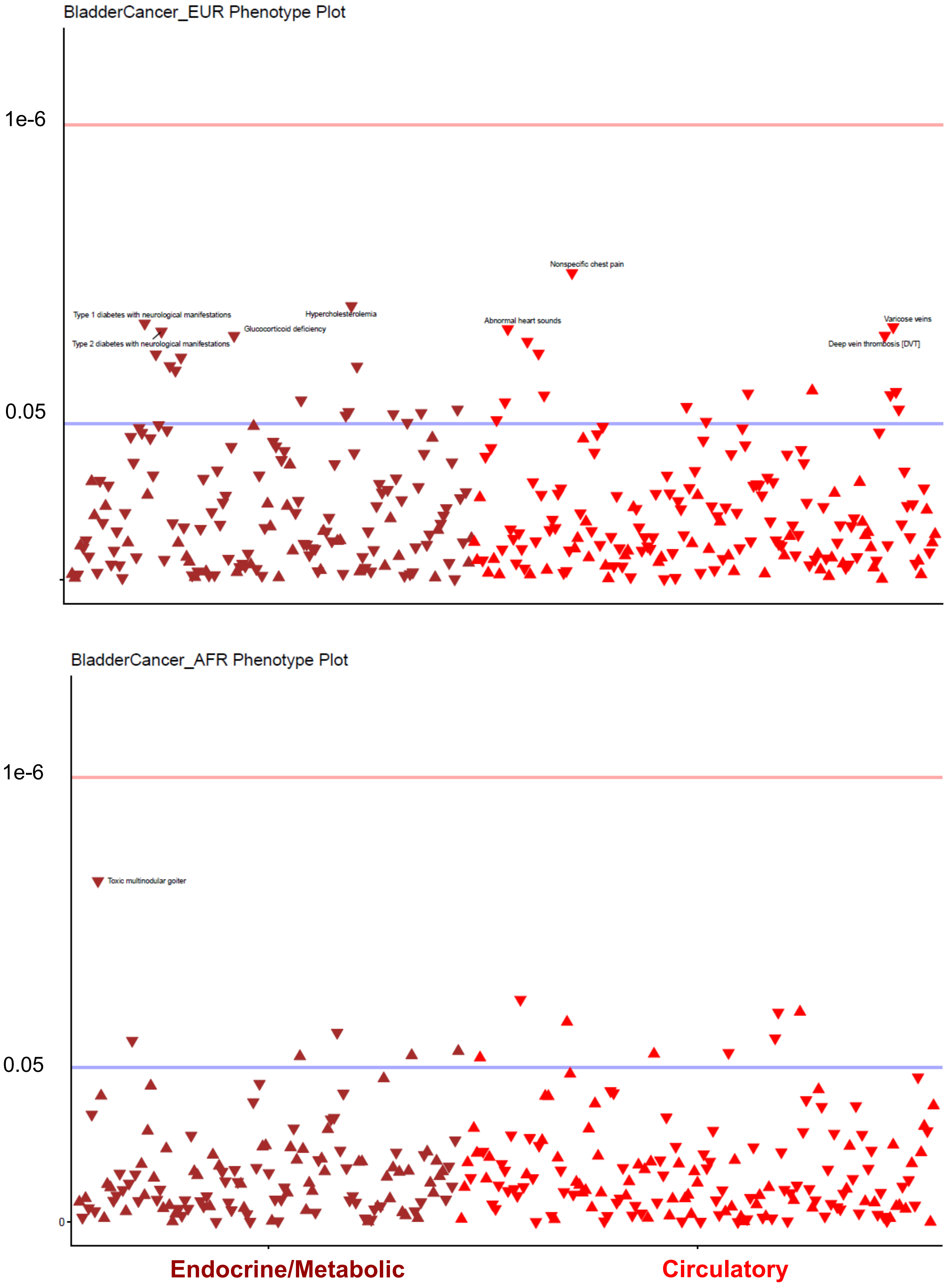


**Supplemental Figure 3: Association of bladder cancer PRS with cardiometabolic phenotypes.** Phewas to examine the association of bladder cancer PRS with circulatory and metabolic/endocrine phecodes in HARE-European and HARE-African individuals.

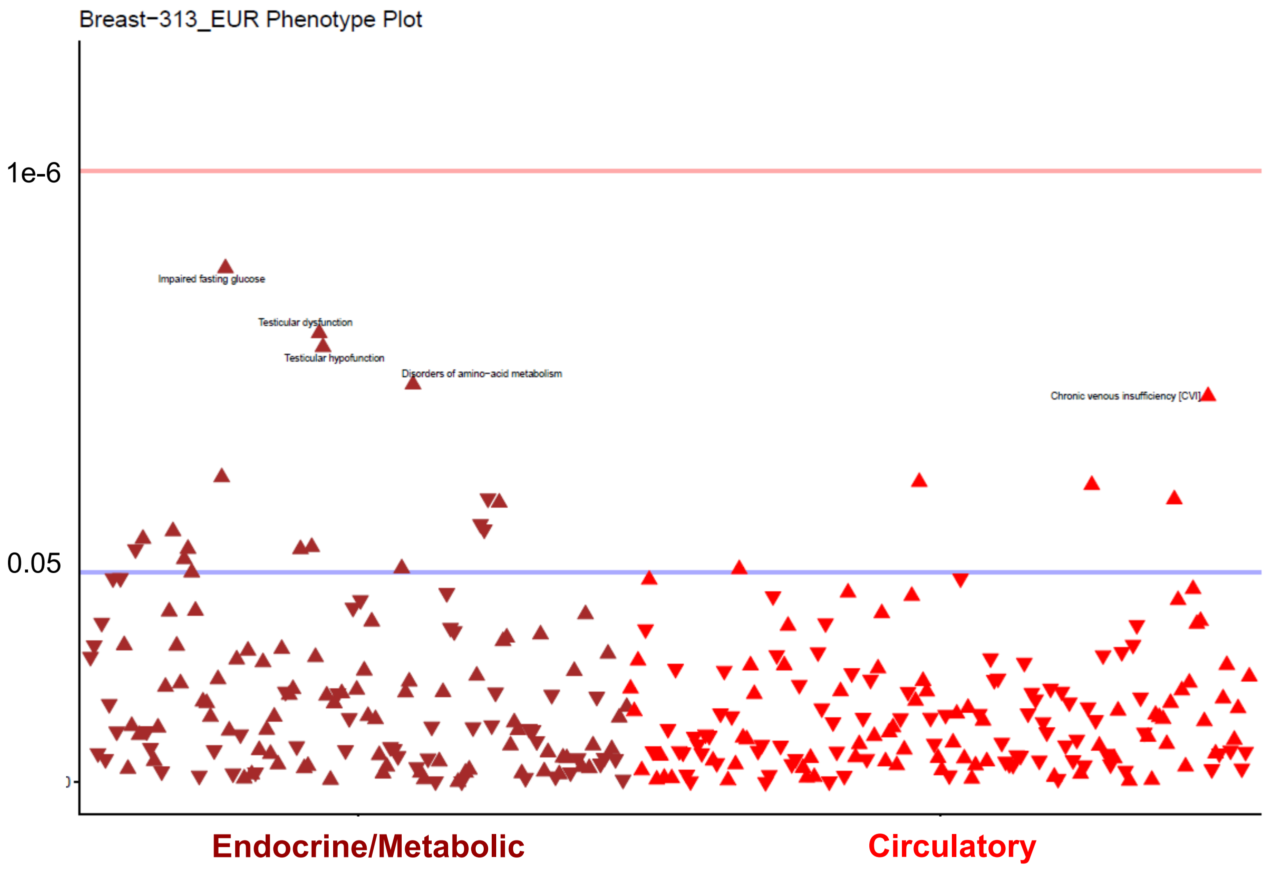


**Supplemental Figure 4: Association of breast cancer PRS with cardiometabolic phenotypes.** Phewas to examine the association of breast cancer PRS with circulatory and metabolic/endocrine phecodes in HARE-European individuals. The breast PRS was not significantly associated with male breast cancer in HARE-African individuals.

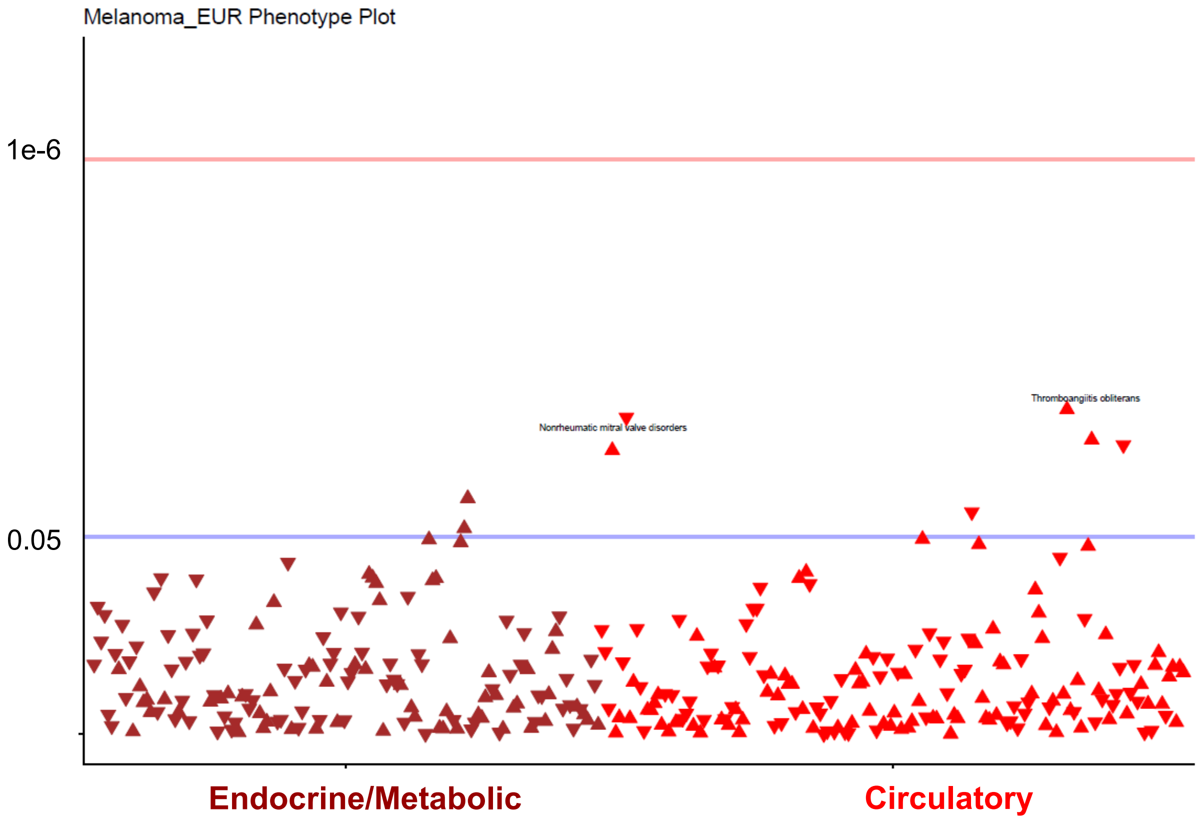


**Supplemental Figure 5: Association of melanoma PRS with cardiometabolic phenotypes.** Phewas to examine the association of melanoma PRS with circulatory and metabolic/endocrine phecodes in HARE-European individuals. The melanoma PRS was not significantly associated with skin cancer in HARE-African individuals.

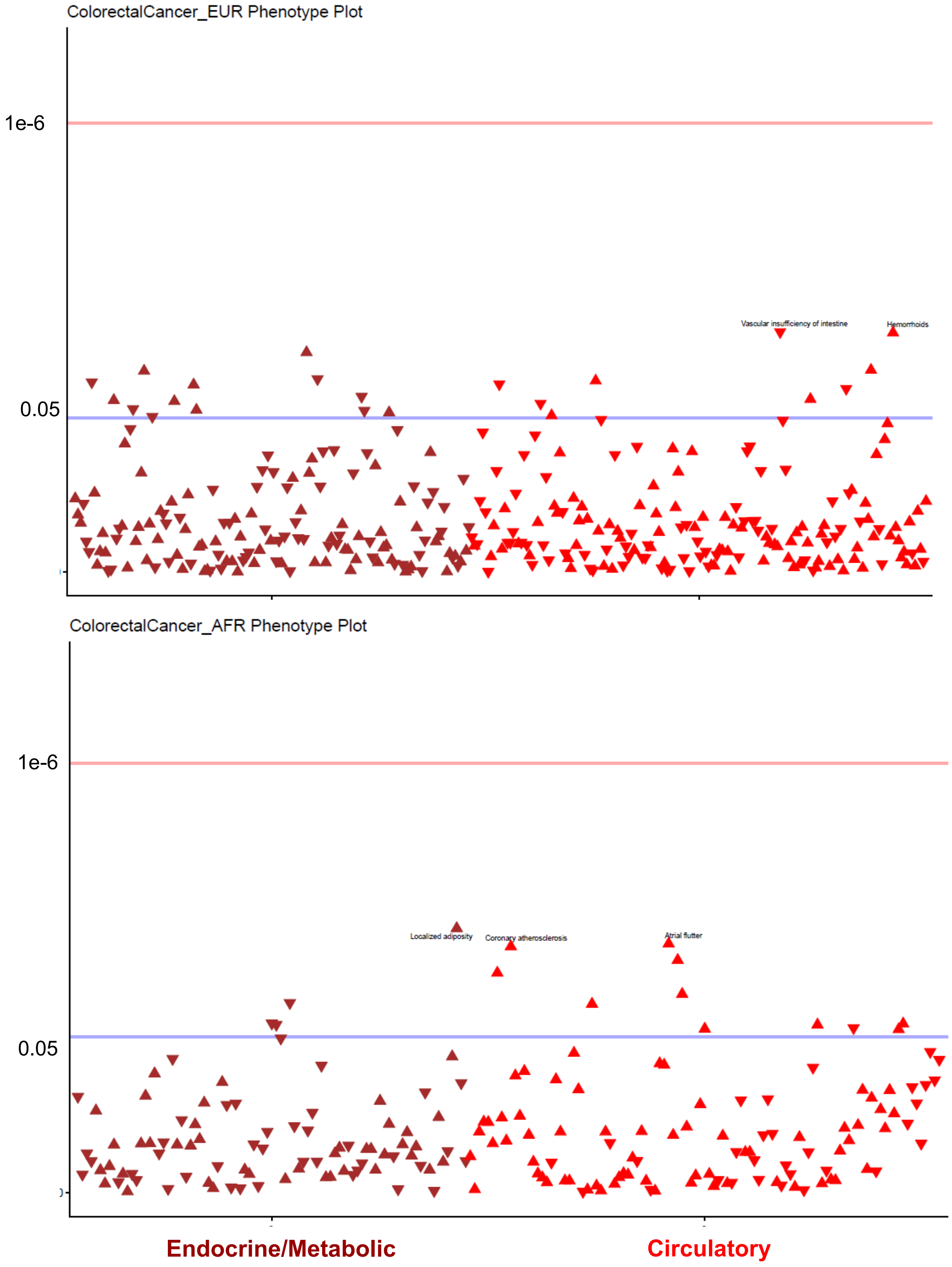


**Supplemental Figure 6: Association of colorectal cancer PRS with cardiometabolic phenotypes. a.** Phewas to examine the association of colorectal cancer PRS with circulatory and metabolic/endocrine phecodes in HARE-European and HARE-African individuals.

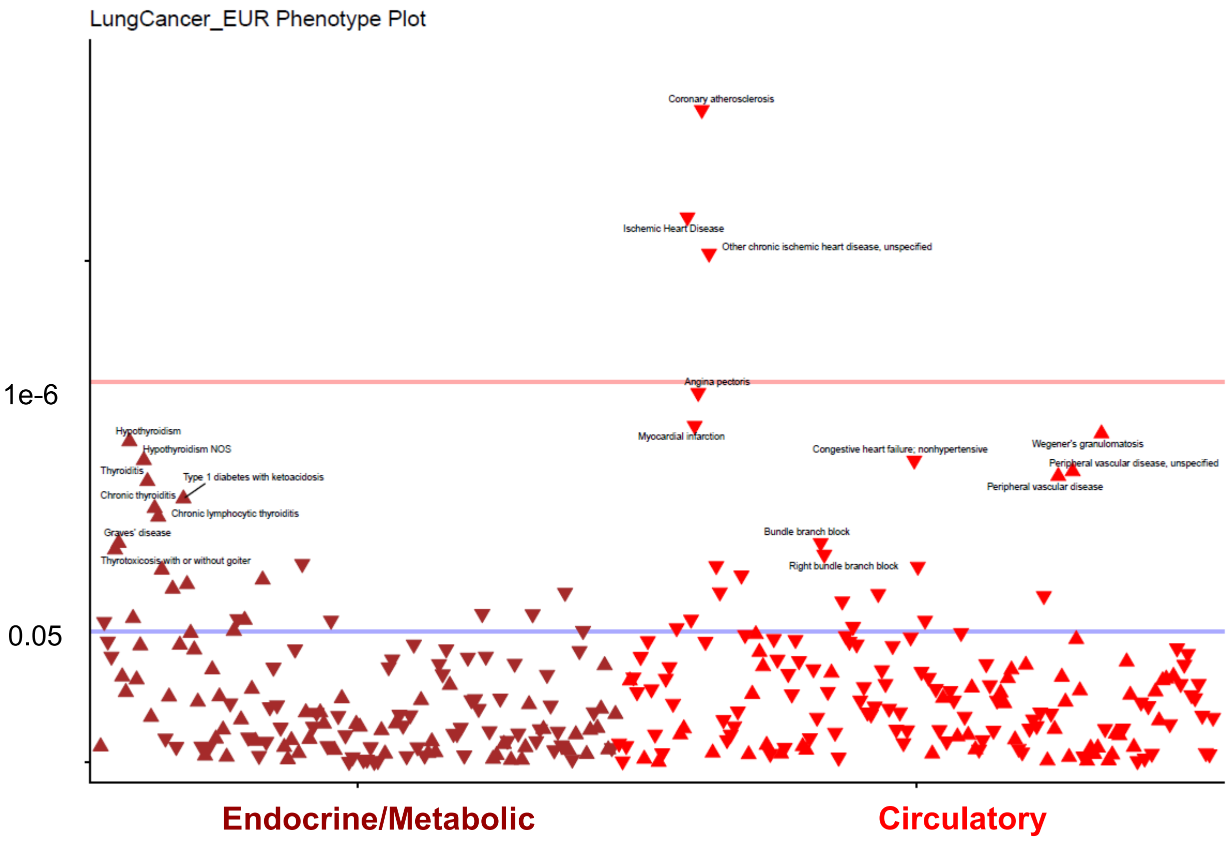


**Supplemental Figure 7: Association of lung cancer PRS with cardiometabolic phenotypes.** Phewas to examine the association of lung cancer PRS with circulatory and metabolic/endocrine phecodes in HARE-European individuals. The lung cancer PRS was not significantly associated with lung cancer in HARE-African individuals.

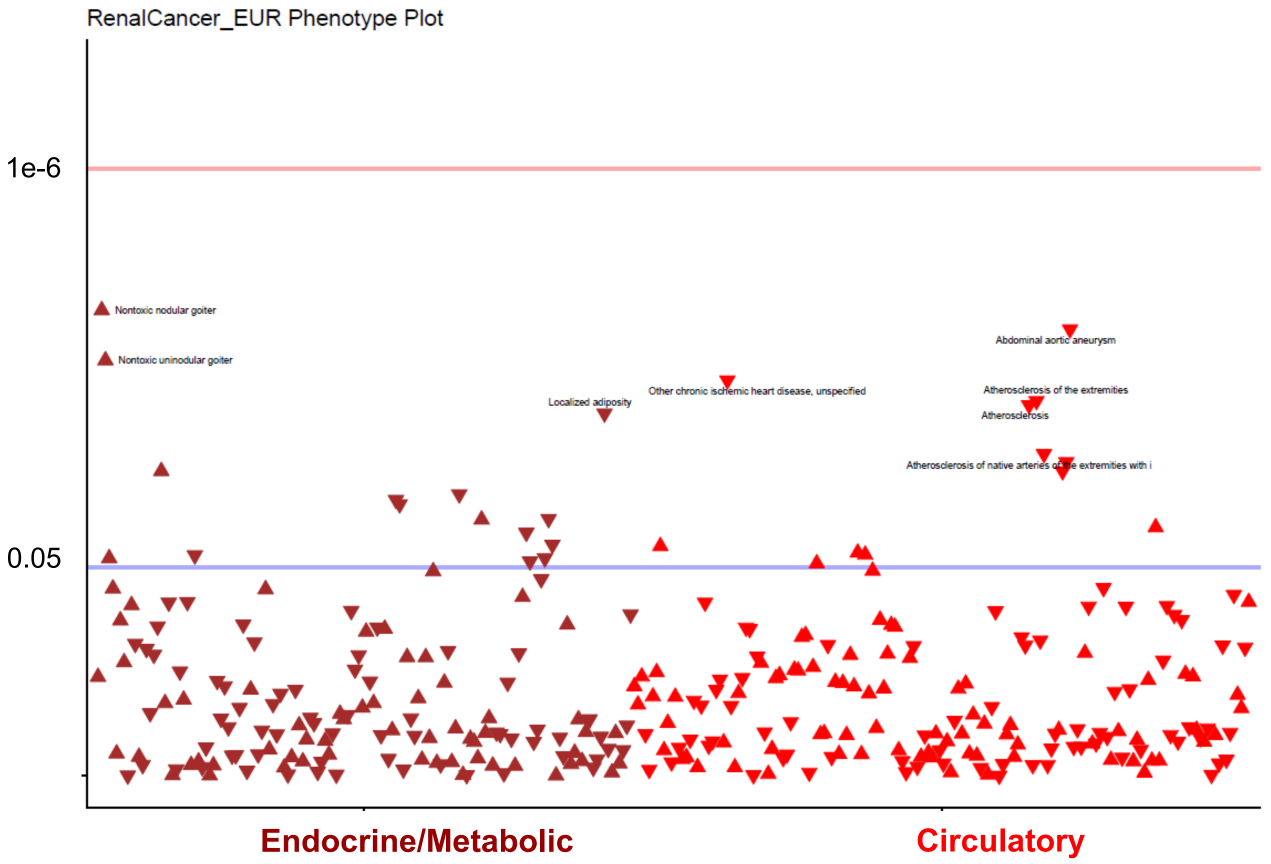


**Supplemental Figure 8: Association of renal cancer PRS with cardiometabolic phenotypes.** Phewas to examine the association of renal cancer PRS with circulatory and metabolic/endocrine phecodes in HARE-European individuals. The renal cancer PRS was not significantly associated with renal cancer in HARE-African individuals.

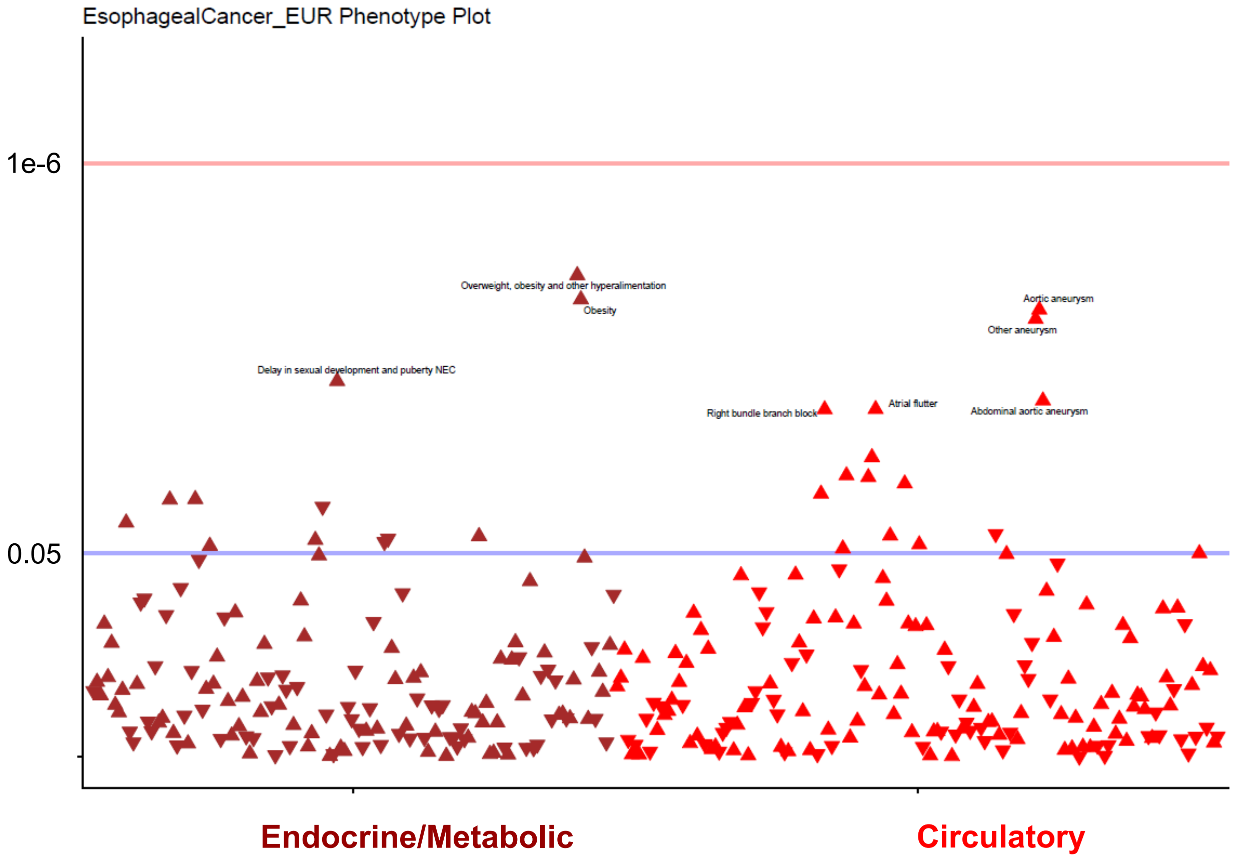


**Supplemental Figure 9: Association of esophageal cancer PRS with cardiometabolic phenotypes.** Phewas to examine the association of esophageal cancer PRS with circulatory and metabolic/endocrine phecodes in HARE-European individuals. The esophageal cancer PRS was not significantly associated with esophageal cancer in HARE-African individuals.

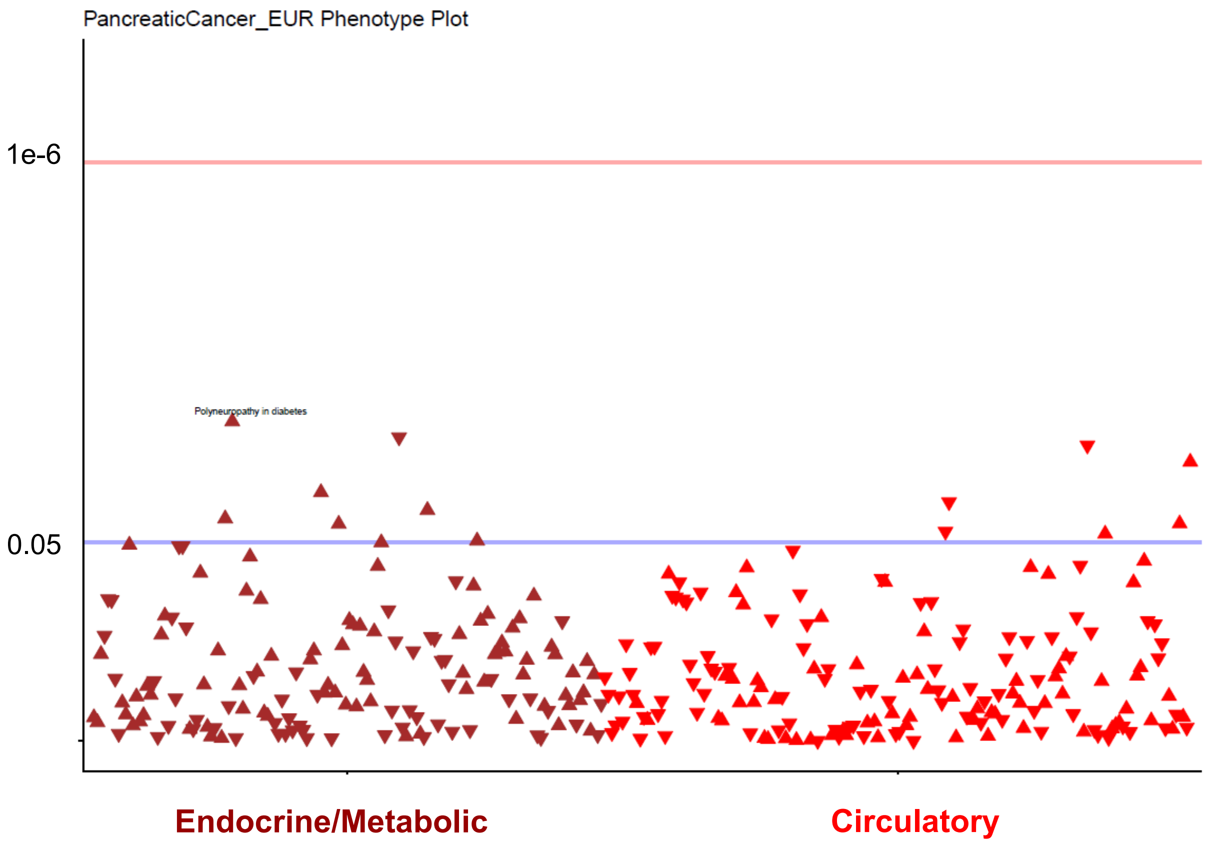


**Supplemental Figure 10: Association of pancreatic cancer PRS with cardiometabolic phenotypes.** Phewas to examine the association of pancreatic cancer PRS with circulatory and metabolic/endocrine phecodes in HARE-European individuals. The pancreatic cancer PRS was not significantly associated with pancreatic cancer in HARE-African individuals.

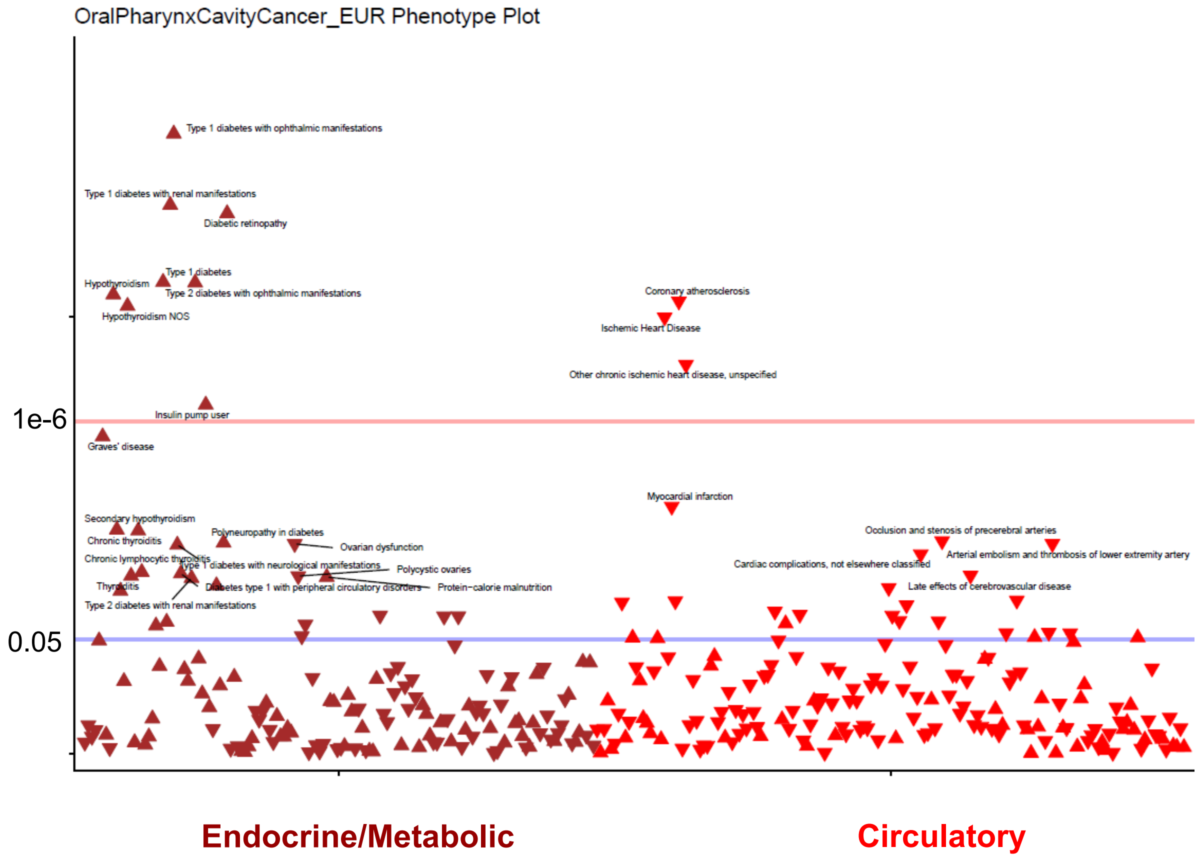


**Supplemental Figure 11: Association of oral cancer PRS with cardiometabolic phenotypes.** Phewas to examine the association of oral cancer PRS with circulatory and metabolic/endocrine phecodes in HARE-European individuals. The oral cancer PRS was not significantly associated with oral cancer in HARE-African individuals.

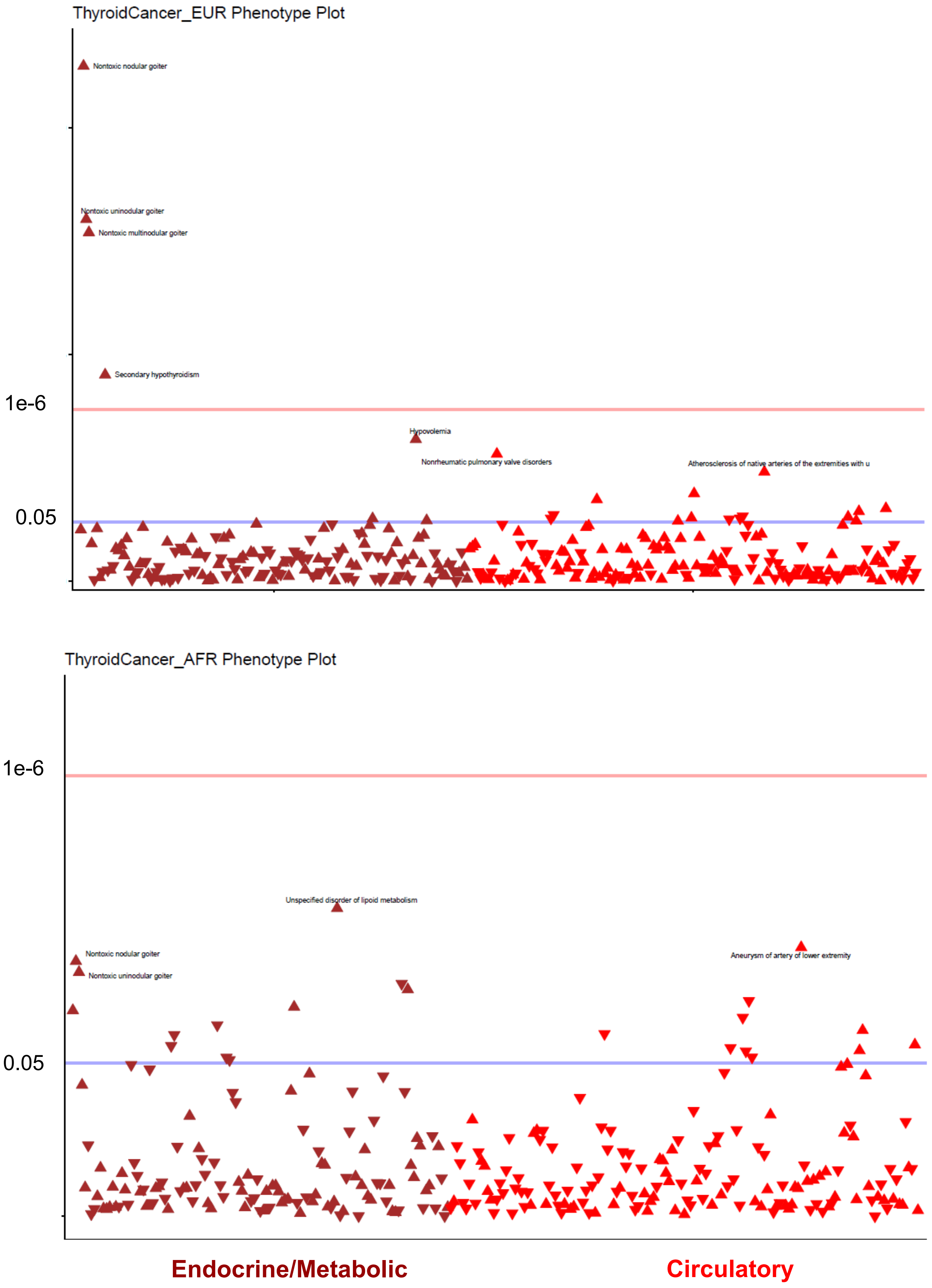


**Supplemental Figure 12: Association of thyroid cancer PRS with cardiometabolic phenotypes. a.** Phewas to examine the association of thyroid cancer PRS with circulatory and metabolic/endocrine phecodes in HARE-European and African individuals.

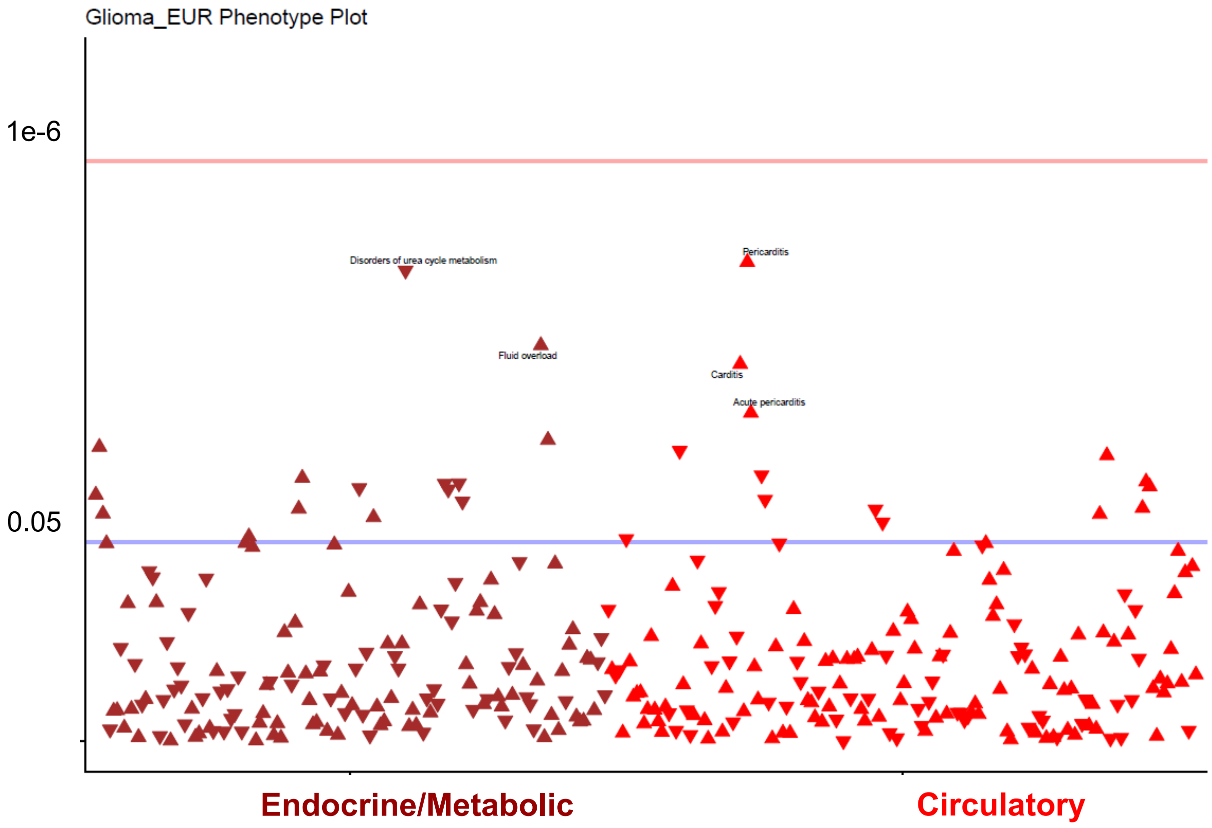


**Supplemental Figure 13: Association of glioma PRS with cardiometabolic phenotypes.** Phewas to examine the association of glioma PRS with circulatory and metabolic/endocrine phecodes in HARE-European individuals. The glioma PRS was not significantly associated with brain cancer in HARE-African individuals.

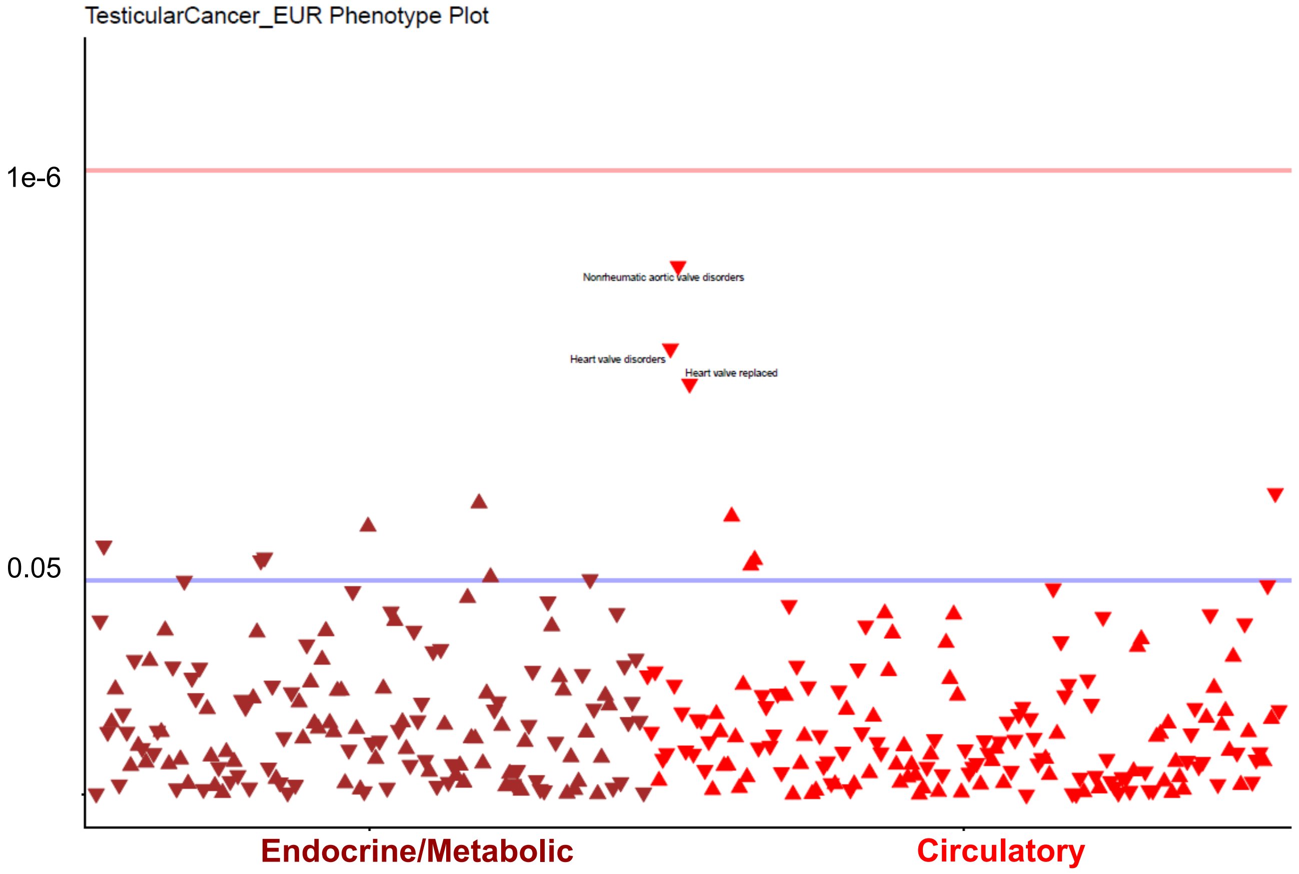


**Supplemental Figure 14: Association of testicular cancer PRS with cardiometabolic phenotypes.** Phewas to examine the association of testicular cancer PRS with circulatory and metabolic/endocrine phecodes in HARE-European individuals. The testicular cancer PRS was not significantly associated with testicular cancer in HARE-African individuals.
